# Detecting multiple sclerosis disease activity and progression in progress notes from electronic medical records using natural language processing and machine learning

**DOI:** 10.1101/2022.10.11.22280951

**Authors:** Jack Z. Chang, Megan H. Hyland, Kathleen Munger, Ryan Canissario, Robert G. Holloway, Jiebo Luo, Timothy D. Dye

## Abstract

Multiple sclerosis (MS) phenotypes provide useful disease descriptions but lack complete information regarding the continuing disease process. Disease activity and progression are meaningful modifiers of the MS phenotypes which can further guide prognosis, therapeutic decisions, and clinical trial designs and outcomes, which were not explicitly documented in patients’ electronic medical records (EMRs). We aimed to detect disease activity and progression in patients with MS from clinical notes in the EMR using Natural Language Processing and Machine Learning models. Using randomly selected progress notes from MS patients at the University of Rochester MS clinic, we integrated NLP and machine learning technologies to predict selected phenotype modifiers that represent disease activity and progression. The method was evaluated by the performance of both the NLP models and machine learning models, as well as the interpretability of the integrated method. We identified 460 progress notes from 287 adult MS patients. The NLP model had an average of 0.92 in precision, 0.87 in recall, and 0.89 in F-score for entity extraction. It had an average of 0.85 in precision, 0.84 in recall, and 0.85 in F-score for entity relation extraction. The sensitivities and specificities of the classification algorithms in predicting phenotype modifiers were: 67% and 93% for predicting modifier “Active”, 61% and 82% for predicting modifier “Worsening”, 92% and 98% for predicting modifier “Progression”, 80% and 94% for predicting modifier “New MRI Lesion”, respectively. We showed that the integrated method of NLP with machine learning classification is capable of detecting evidence of disease activity and clinical progression from clinical notes. The classification algorithms yielded interpretable and largely clinically relevant features (symptoms and clinical conditions) that were persistently associated with disease activity and progression. This method holds promise for facilitating the screening of MS clinical trial participants and potentially identifying early evidence of disease progression.

**Author Summary:** Disease activity and progression of disability can be meaningful modifiers to base MS phenotypes which can further impact prognosis, therapeutic decisions, and clinical trial designs and outcomes. However, studies have shown that neither MS phenotypes nor their modifiers are consistently documented in electronic medical record (EMR) chart notes. The evidence for disease activity and progression often resides in the clinical notes, requiring manual chart review from clinical experts and increasing the difficulty of conducting clinical research. In this paper, we developed a generalized information extraction, classification and prediction pipeline, incorporating Natural Language Processing (NLP) technologies and shallow machine learning models, to detect MS disease activity and progression in clinical notes from EMR and to predict phenotype modifiers. Results demonstrated that this integrated method extracts clinically relevant information from progress notes that are persistently associated with disease activity and progression, and predicts MS phenotype modifiers with satisfactory performance, encouraging portability and interpretability. In the future, we aimed to apply the method in this study for facilitating high throughputs of MS clinical trial screening and assessing disease modifying therapy utilization based on disease modifiers.

## 1. Introduction

Multiple sclerosis (MS) is an inflammatory disorder where the body’s immune system targets the central nervous system (CNS), causing disrupted nerve signals through demyelination and axonal degeneration. MS is the most common cause of nontraumatic neurologic disability in young adults [1], affecting around 900,000 people in the United States [2]. MS is associated with significant costs, disability, and decreased quality of life for patients and their families [3, 4].

MS is characterized into different clinical courses or phenotypes, including clinically isolated syndrome, relapsing-remitting, secondary progressive, and primary progressive. These existing MS phenotypes can be useful for standardizing communication about patients, selecting appropriate therapies, and identifying clinical trial candidates. The clinical phenotype may be assessed based on current status and historical data, with the understanding that MS is a dynamic process and that the initially assessed phenotype may change over time. Patients with a relapsing-remitting course may transition to develop more secondary progressive features. Conversely, patients with a progressive course may have evidence of relapsing activity, including acute attacks and/or new asymptomatic contrast-enhancing brain lesions, particularly earlier in the disease. Accurate clinical MS phenotypes as the disease course evolves are critical for individualized clinical and research decision-making [5]. However, the clinical course is quite variable, and the phenotypes alone may not fully describe the continuing disease process. In 2011, the International Advisory Committee on Clinical Trials of MS and the MS Phenotype Group re-examined the core MS phenotypes and recommended modifiers of these phenotypes, including the assessment of disease activity, as defined by clinical assessment of relapse occurrence or lesion activity, and determination of whether progression of disability has occurred [5]. These additional disease modifiers may aid physicians with prognostication and treatment adjustments to improve quality of life and slow disability progression.

Neither MS phenotypes nor modifiers are consistently documented in electronic medical record (EMR) chart notes [6]. The administrative coding of MS is wholly contained as a single diagnosis code (G35.0 in ICD-10-CM or 340.0 in ICD-9-CM) that is uninformative about MS phenotypes or modifiers. There are no blood or cerebrospinal fluid (CSF) biological markers to date that can reliably and reproducibly differentiate between MS clinical phenotypes [5]. The evidence for disease activity and progression often resides in the clinical notes in the EMR even if not explicitly documented. Therefore, extensive chart review and abstraction from the EMR is often needed to collect phenotypes and modifiers information on MS patients for activities such as clinical trials and observational studies, thus increasing the difficulty of conducting clinical research.

Researchers have applied natural language processing (NLP) techniques on clinical text in the EMR to identify MS phenotypes [6-9]. The popular approaches involve interviewing clinical experts for possible keywords and phrases denoting MS phenotype in order to develop a data dictionary, applying NLP methods to perform text search, and using the search outcomes (i.e., mentions of the dictionary terms) as input to either prediction models or rule-based algorithms. Nelson et al. developed possible keywords and phrases denoting MS phenotype contributed by clinical experts (a registered nurse, a physician therapist, and a licensed family counselor), and applied NLP to conduct a keyword search in clinical notes with negation detection [6]. Davis et al. applied keyword search and regular expression to extract four clinical subtypes from clinical notes, letters and problem lists that mentioned MS [8]. Xia et al. generated a list of clinician expert-defined, MS-relevant codified and narrative variables from the EMR data for each patient and extracted variables with 10% or more frequency of occurrence [10]. These approaches either aimed to develop an algorithm for classifying the disease (MS) itself (not the phenotype), or to determine the phenotype prevalence in clinical notes in a large patient population. A limitation of their works was the lack of focus on determining the MS phenotypes and/or their modifiers. Additionally, the approaches demonstrated in those studies required a task-specific dictionary of relevant phrases or a set of hand-crafted rules [11]. The development of a dictionary and rules usually requires significant effort and an understanding of the task from domain experts. A rule-based application may not be portable beyond the use case for which it is designed [12]. Lastly, the prediction models are challenging to interpret, and the applied NLP methods are relatively rudimentary and can be further improved.

The objective of this study was to detect MS phenotype modifiers (i.e., disease activity and progression) in clinical notes from the EMR using Natural Language Processing (NLP) and Machine Learning models. To our knowledge, this is the first study to make use of NLP and machine learning technologies to predict these “real-time” modifiers of MS phenotypes in a large patient population.

## 2. Methods

### 2.1 General Approach

We integrated NLP and machine learning technologies to develop our generalized classification and prediction pipelines (Figure 1). A study corpus was created by randomly selecting progress notes in study subjects’ EMRs from their office visits during the study time window. We identified four MS phenotype modifiers, including “Active”, “Worsening”, “Progression”, and “New MRI Lesion”, to represent disease activity and progression. Each note in the study corpus was manually labeled with all four modifiers using a binary class. The natural language processing pipeline was developed to extract symptoms and clinical conditions from the study corpus that could serve as evidence of MS disease activity and progression during the assessment period. These NLP extractions were normalized to the standard controlled vocabulary of Unified Medical Language System (UMLS) from US National Library of Medicine (NLM), and were subsequently used as input features to train supervised machine learning classification models to predict those MS phenotype modifiers. The method was evaluated by the performance of both the NLP models and machine learning models, as well as the interpretability of the integrated method.

**Figure 1.**
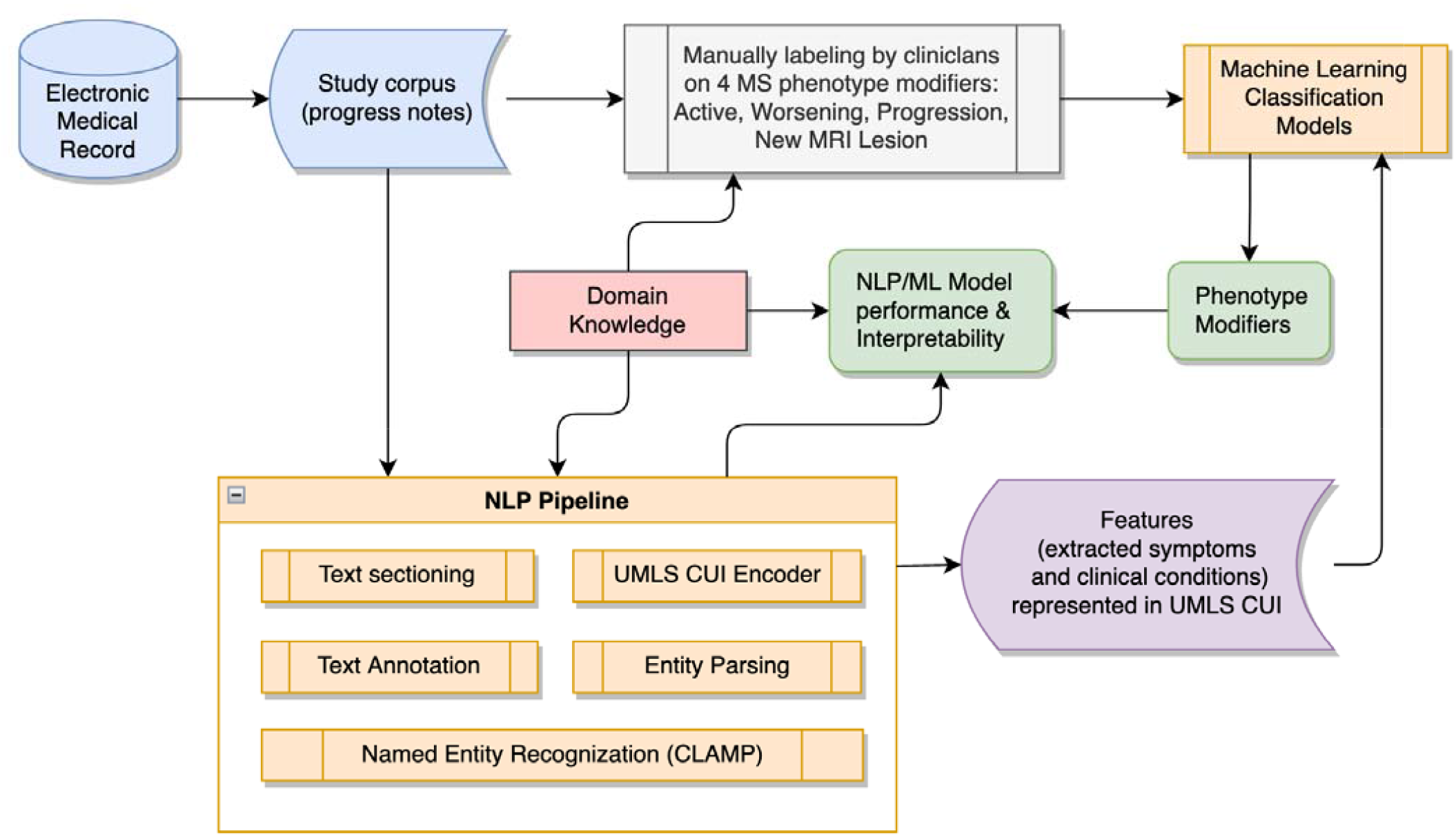
Integrated NLP and machine learning pipelines to predict MS phenotype modifiers for MS disease activity and progression

### 2.2 Patient Selection

This study used EMR data from the MS clinic at the University of Rochester Medical Center to identify candidate patients with at least one diagnosis of MS (ICD-10-CM code G35.0 or ICD-9-CM code 340.0) who were over 18 years old and received care from the MS clinic and had at least one office visit between January 1 2016 and December 1 2020. The selection of office visits was limited to the primary MS clinic providers which included both physicians (attendings and fellows) and advanced practice providers. Progress notes were collected from the completed office visits of candidate patients. Study corpus was then created by randomly selecting candidate patients and their progress notes which were stratified by visit year and provider.

### 2.3 Progress Notes Labeling

To quantify disease activity and progression during the assessment period, three MS-trained physicians formed a review group and defined four MS phenotype modifiers to represent disease activity and progression, including “Active”, “Worsening”, “Progression”, and “New MRI Lesion”. “New MRI Lesion” was marked “true” for MRI activity, defined as a contrast-enhancing lesion and/or one or more new or unequivocally enlarging T2 lesions. “Active” was marked “true” for either a clinical relapse (new or significantly worsened symptoms lasting >24 hours in the absence of other obvious precipitating factors such as underlying infection) and/or MRI activity as defined above. “Progression” was marked “true” if the note indicated gradual worsening of symptoms by history or objective worsening on exam noted by the provider. “Worsening” was a composite marker indicating either activity or progression; it was marked “true” if any of the other 3 modifiers were true.

In some instances, activity was acutely detailed in a progress note (i.e., a recent MRI with a contrast-enhancing lesion). In other cases, the activity or progression was a relative measure compared with the last progress note (i.e., a new MRI lesion compared with 6 months prior). To account for this, short interval follow-up notes (i.e., < 6 months apart) were still labeled “Active” in the setting of recent relapse, even if not “new” compared to prior follow up. Conversely, “Progression” or “New MRI Lesion” may not have been marked “true” if the comparison note was remote or unclear. To account for this variability as well as the subjectivity in classifying relapses/progression, multiple clinicians reviewed the progress notes. The first 150 progress notes were reviewed by all three clinicians and then reviewed as a group for consensus. Subsequent notes were reviewed by a single clinician, but any potential discrepancies were flagged and then reviewed again by the group for consensus. Additionally, notes that were mislabeled as MS (i.e., patients with other neuroimmunologic diseases) or notes that were not true progress notes (i.e., therapy monitoring appointments) were excluded. A total of 21.4% of notes were excluded from the study corpus.

### 2.4 Natural Language Processing (NLP) pipeline

To assess disease activity and to determine disease progression, we extracted MS-related symptoms and clinical conditions from progress notes which were either newly developed since the last visit or represented the disease progression during the visit intervals. The extractions were used as input features in the subsequent machine learning classifier to predict phenotype modifiers. The appropriate clinical feature representation has been shown to improve the performance of machine learning classifiers [13]. To do this, we adopted an NLP toolkit Clinical Language Annotation, Modeling, and Processing (CLAMP) [14], and used the UMLS Metathesaurus to normalize clinically-relevant UMLS concepts in progress notes.

The outpatient progress notes are parts of EMRs where healthcare providers document patient’s clinical status over the course of an outpatient visit. These notes do not always have a consistent template or documentation pattern and usually consist of many sections, including Disease Summary, History of Present Illness, Interval History, Past Medical History, Current Outpatient Prescriptions, Review of Systems, Physical Exam, Assessment, and Plan. Each section may start with a section header, but the wording of the section header and position of each section within progress notes was not always consistent. The evidence of disease activity and progression was available in the progress note sections of Interval History, HPI, Subjective, and MS History, presented here in the order of precedence. We analyzed the documentation patterns and developed regular expression algorithms to section the progress notes, and removed the un-relevant sections in each progress note as we were only interested in the part of progress notes that represented the disease activity and progression during the assessment period.

The NLP process we built in CLAMP included the following major modules: Sentence detector, Tokenizer, POS tagger, Chunker, Named Entity Recognizer, Assertion and negation. We developed annotation guidelines, annotated progress notes with 8 entities and 7 entity relations (see Table 1), and re-trained the Named Entity Reorganization (NER) model with 5-fold cross-validation.

**Table 1.** Entity and Entity Relations in Progress Note Annotation

|  |  |  |  |
| --- | --- | --- | --- |
| **Semantic**<br>**: Entity** | **Semantic Description** | **Semantic :**<br>**Entity** | **Semantic**<br>**Description** |

Table 1. Entity and Entity Relations in Progress Note Annotation
|  |  | Relations |  |
| --- | --- | --- | --- |
| Problem | symptoms, such as “enhancing lesion”, “stiffness/spasticity in the lower extremities”, “urinary urgency”, “new neurologic symptoms”, etc. | -- | -- |
| NEG | negation, such as “denies”, “did not have”, “no evidence of”, etc. | NEG_Of | negation of the problem |
| SEV | severity, such as “mild”, “severe”, “acute”, etc. | SEV_Of | severity of the problem |
| BDL | body location such as “both legs”, “Rt knee”, etc. | BDL_Of | body location of the problem |
| SUB | subject, such as “her mother”, “her aunt”, etc. | SUB_Of | subject of the problem |
| COU | course, such as “worsening”, “improved”, “resolved”, etc. | COU_Of | course of the problem |
| UNC | uncertainty, such as “does not recall”, “may have”, etc. | UNC_Of | uncertainty of the problem |
| temporal | temporal expression, such as “in 2016”, “the fall of 2012”, “12/01/16”, etc. | hasTemporal | temporal expression of the problem |

The NER model extracted symptoms and clinical conditions that represented the patient’s clinical status during the assessment period. As previously noted, we identified only those that were either newly developed since the last visit or represented the disease progression during the visit intervals. We examined NER extractions and developed Entity Parser to parse the NER outputs and to exclude the non-eligible symptoms. The symptoms were considered not eligible if they were negated, or were uncertain, or were related to other family members rather than patients themselves, or were reported as part of medical history, or were resolved, gone, or improved in the assessment period.

Extracted symptoms often consist of multiple words and have variations of how they are worded and documented by different providers or by the same provider at different time. For example, the following phrases have the same semantic interpretation but appeared equally in the progress notes: “poor short-term memory”, “short term memory issues”, “short-term memory difficulty”, “mild difficulty short-term memory”. After identifying the eligible symptoms, they were then encoded to the controlled biomedical vocabulary of NLM’s UMLS. Each term in the UMLS metathesaurus is presented by a unique concept identifier (CUI). The abovementioned four-symptom phrases were encoded and normalized to a single UMLS concept “Poor short-term memory” with CUI C0701811.

### 2.5 Machine Learning Model Development

The extracted symptoms and clinical conditions from the NLP pipeline were normalized and encoded to standard UMLS CUI. The CUIs were transformed to dichotomous CUI variables using one-hot encoding. Six supervised machine learning models with feature selection were developed to predict those four MS phenotype modifiers individually, using the dichotomous CUI variables as input features and MS phenotype modifiers as outcomes. The machine learning models included L1-regulated logistic regression (LASSO), regularized SVM with linear kernel, random forest, decision tree, linear Discriminant Analysis (LDA), and Naïve Bayes (NB). Five-fold cross-validation was applied in each model to reduce overfitting and to evaluate model performance. Since all of the modeling processes were developed independently for every phenotype modifier, each model is a binary classifier instead of multi-class classifier, which reduces the evaluation complexity [13].

### 2.6 Evaluation

The evaluation consisted of three parts. Firstly, the NLP pipeline was evaluated on the notes comprising the test set. The primary outcome of the NLP pipeline was the performance of abstracting entities and entity relations from notes. We calculated Precision, Recall and the F1 score for each one of entities and entity relations compared with the reference standard, which was the manually annotated entities and entity relations in the test set. Precision is the ratio of correctly predicted positive observations to the total predicted positive observations, whereas recall is the ratio of correctly predicted observations to the all actually positive observations. Precision represents what percent of entities or entity relations predication were correct. Recall represents what percent of the real entities or entity relations the NLP model caught. The F1 score is the weighted average of Precision and Recall. This score takes both false positives and false negatives into account. It is the harmonic mean of the both Precision and Recall. Secondly, the machine learning classification models were evaluated by their performance of predicting the MS phenotype modifiers. We determined the sensitivity and specificity of each model and modifier compared with the reference standard, which was the manually labeled modifiers on progress notes. To determine sensitivity and specificity, each modifier is considered as a binary class, so that we obtained true positive, false positive, true negative, and false negative for each class. Finally, we evaluated the interpretability by selecting input features with high coefficients and discussed how relevant they were in clinical settings to assess disease activity and to determine disease progression.

### 2.7 Ethics Statement

All relevant ethical safeguards have been met in relation to patient privacy protection. Institutional review board (IRB) approval for this study was obtained through the University of Rochester’s Research Subjects Review Board (RSRB) under the Office of Human Subject Research. The approval number is STUDY00005629. The RSRB granted waiver of Informed Consent and waiver of HIPAA authorization to this study because the research involves medical record review only and is no greater than minimal risk and there is no recruitment or intervention performed.

## 3. Results

### 3.1 Dataset and Study Corpus

A total of 1,265 patients with MS diagnoses had visits listed at the UR MS clinic with office visits between January 1 2016 and December 1 2020. “No Show” or “Canceled” office visits were excluded. A total of 4,095 office visits were identified in which neurology progress notes were available and documented by the selected types of clinicians. Visits with other providers (such as nurses, therapists) were excluded because the clinical notes from those visits did not always have complete information on patients’ clinical status. The study corpus included 460 progress notes from 287 MS patients which were randomly selected from the candidate patient cohort, stratified on visit year and provider, to maximize the independence among the observations. Each progress note was manually labeled on those four phenotype modifiers. The demographic characteristics of study subjects and study corpus are listed in Table 2a and Table 2b.

**Table 2a.** Demographic characteristics of study subjects

| Category | Items | Patient Count |
| --- | --- | --- |
| Total # of patients |  | 287 |
| Gender | Female | 205 (71.4%) |
|  | Male | 82 (28.6%) |
| Race | Black or African American | 28 (9.8%) |
|  | White | 236 (82.2%) |
|  | Other / Unknown / Patient Refused | 23 (8.0%) |
| Ethnicity | Hispanic or Latino | 12 (4.2%) |
|  | Not Hispanic or Latino | 228 (79.4%) |
|  | Unknown | 47 (16.4%) |
| Age at office visit | 18-20 | 2 (0.7%) |
|  | 21-40 | 100 (34.8%) |
|  | 41-60 | 144 (50.2%) |
|  | 61-80 | 43 (15.0%) |
|  | 80+ | 1 (0.3%) |

**Table 2b.** Distribution of phenotype modifiers in study corpus

| Label | Modifier "Active" | Modifier "Worsening" | Modifier "Progression" | Modifier "New MRI Lesion" |
| --- | --- | --- | --- | --- |
| True | 164 | 218 | 61 | 116 |
| False | 292 | 238 | 398 | 240 |
| N/A | 4 | 4 | 1 | 104 |

### 3.2 NLP Pipeline including Text Sectioning, Annotation, NER, Entity Parsing, and CUI Encoder

Progress notes in the study corpus were sectioned and only have the section(s) of Interval History, HPI, Subjective, and MS History retained. They were then annotated individually with study defined 8 entities and 7 entity relations (Figure 2) using CLAMP.

**Figure 2.**
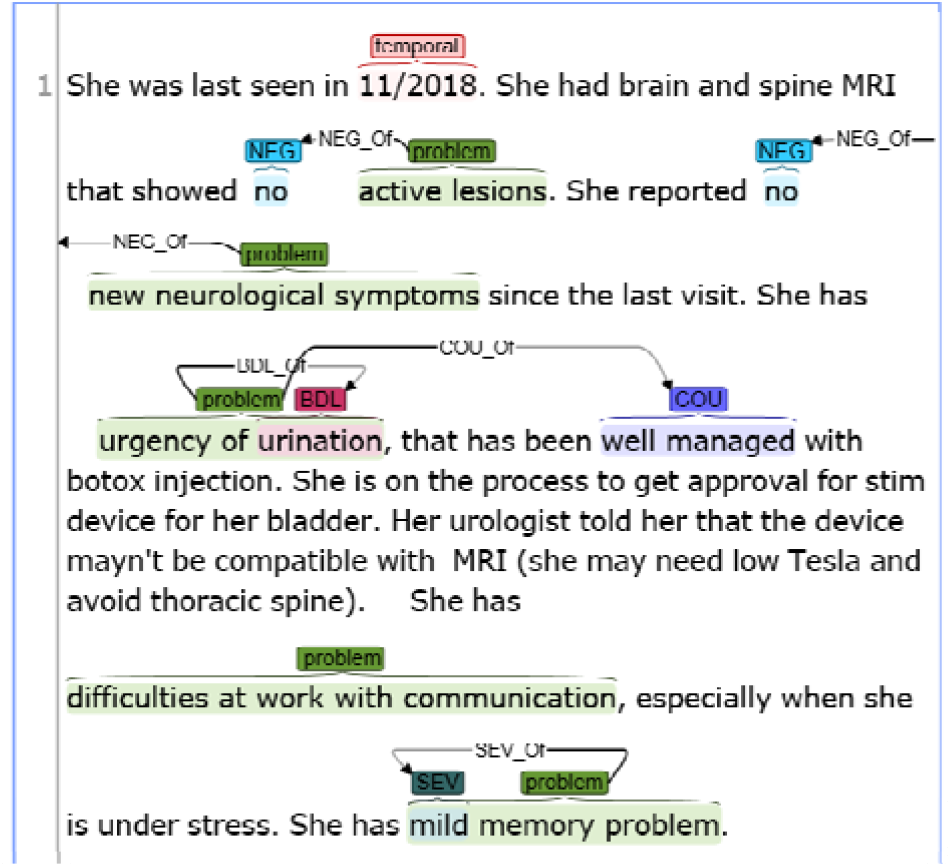
MS progress note annotation with study defined entities and entity relations using CLAMP

After re-training the NLP model with 5-fold cross-validation on annotated notes, a total of 4726 semantic entities and 2331 entity relations were extracted from the study corpus. The NLP model had an average of 0.92 in precision, 0.87 in recall, and 0.89 in F-score for entity extraction. The model had an average of 0.85 in precision, 0.84 in recall, and 0.85 in F-score for (entity) relation extraction. Breakdown of model performance on individual entity and entity relation are listed in Table 3a and Table 3b.

**Table 3a.** NLP performance on entity extraction

| Semantic | Precision | Recall | F1 score |
| --- | --- | --- | --- |
| Problem | 0.805 | 0.701 | 0.750 |
| NEG | 0.983 | 0.987 | 0.985 |
| SUB | 0.833 | 0.833 | 0.833 |
| SEV | 1.000 | 0.818 | 0.900 |
| COU | 1.000 | 0.991 | 0.996 |
| BDL | 1.000 | 0.983 | 0.991 |
| UNC | 1.000 | 0.971 | 0.986 |
| temporal | 1.000 | 1.000 | 1.000 |
| Overall entity extraction | 0.921 | 0.866 | 0.892 |

**Table 3b.**
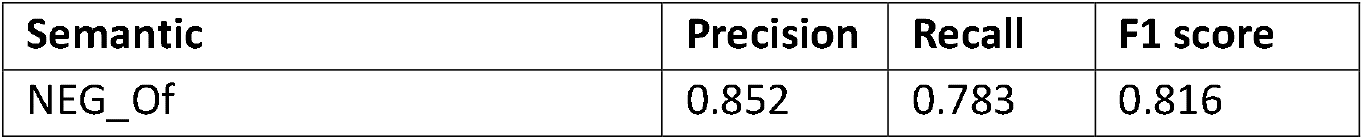
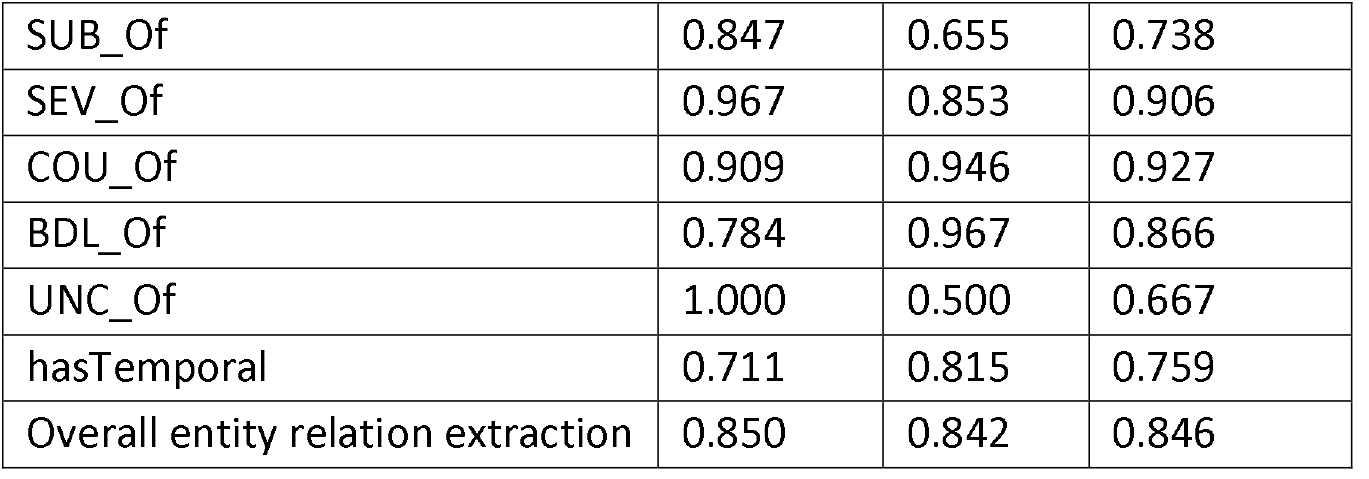
NLP performance on entity relation extraction

The extracted entities and entity relations were run through the Entity Parser to identify eligible symptoms. Extracted entities were considered not eligible symptoms if they were negated, or were uncertain, or were related to other family members rather than patients themselves, or were reported as part of medical history, or were resolved, gone, or improved in the assessment period. Out of extracted semantic entities from the notes, 2404 were eligible symptoms for representing as evidence of the MS disease activity and/or progression over the assessment period. These symptoms were encoded and normalized to 787 unique UMLS CUIs, which were then transformed to dichotomous variables.

### 3.3 Machine Learning Classification and Algorithm Interpretability

Four datasets were built using the 787 binary CUI variables and each one of those 4 MS phenotype modifiers. Each dataset was a 460×788 sparse matrix, with one progress note per row. Each dataset was randomly split to the training (70%) and testing (30%) data sets. Six shallow machine learning classification models were trained using the training dataset and validated on the testing dataset. The best performance of sensitivity and specificity was achieved with L1-regulated logistic regression (LASSO) with feature selection (Table 4). The sensitivities and specificities of the classification algorithms were: 67% and 93% for predicting modifier “Active”, 61% and 82% for predicting modifier “Worsening”, 92% and 98% for predicting modifier “Progression”, 80% and 94% for predicting modifier “New MRI Lesion”, respectively.

**Table 4.** Machine learning classification performance for MS phenotype modifiers

| Model | Modifier<br>“Active” |  | Modifier<br>“Worsening” |  | Modifier<br>“Progression” |  | Modifier<br>“New MRI Lesion” |  |
| --- | --- | --- | --- | --- | --- | --- | --- | --- |
|  | Sensitivity | Specificity | Sensitivity | Specificity | Sensitivity | Specificity | Sensitivity | Specificity |
| Logistic Regression – Lasso | **0.67** | **0.93** | **0.61** | **0.82** | **0.92** | **0.98** | **0.80** | **0.94** |
| Random Forest | 0.67 | 0.89 | 0.62 | 0.77 | 0.91 | 0.97 | 0.76 | 0.88 |
| SVM (Linear) | 0.70 | 0.87 | 0.62 | 0.74 | 0.86 | 0.93 | 0.75 | 0.88 |
| Decision Tree | 0.68 | 0.71 | 0.60 | 0.66 | 0.80 | 0.86 | 0.73 | 0.83 |
| Linear Discriminant Analysis | 0.64 | 0.78 | 0.55 | 0.71 | 0.86 | 0.92 | 0.73 | 0.86 |
| Naïve Bayes | 0.61 | 0.75 | 0.55 | 0.78 | 0.43 | 0.40 | 0.80 | 0.96 |

To evaluate the algorithm interpretability, we further examined the selected input features (i.e., CUI variables representing the MS symptom and clinical conditions) and their coefficients out of the LASSO regression model. Since the datasets are all sparse, the LASSO regression model performs feature selection by adding penalization to weak predictors, hence eliminating them from the model. The coefficient represented how strongly each selected predictor (i.e., CUI variable) is correlated with the prediction outcome (i.e., the phenotype modifiers). Since the coefficients can’t be compared across models, we ranked them by their absolute values to represent how CUI variables were correlated with individual modifier and were associated among multiple modifiers. Higher absolute values of coefficients were associated with lower rank. The top selected input features with highest absolute value of coefficients and their ranking within each MS phenotype modifier were listed in Table 5.

**Table 5.**
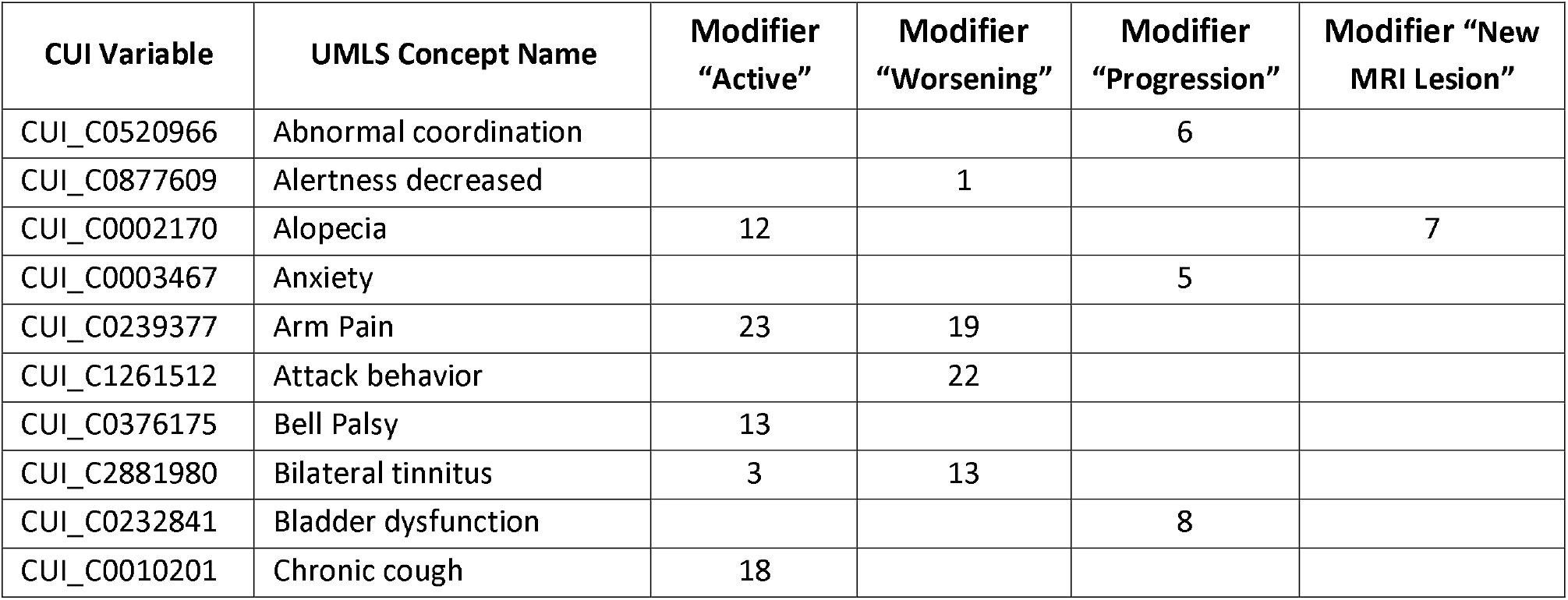
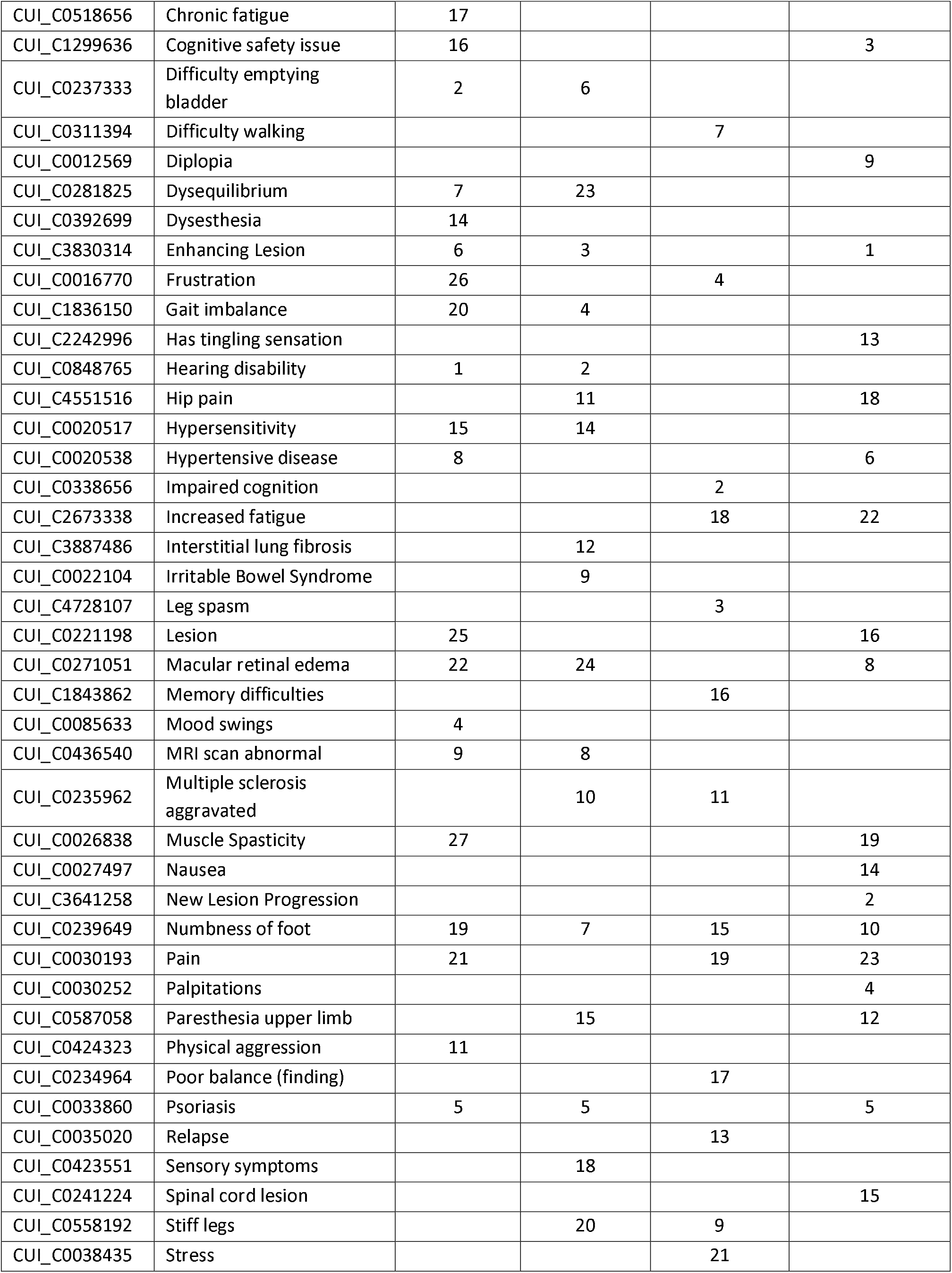
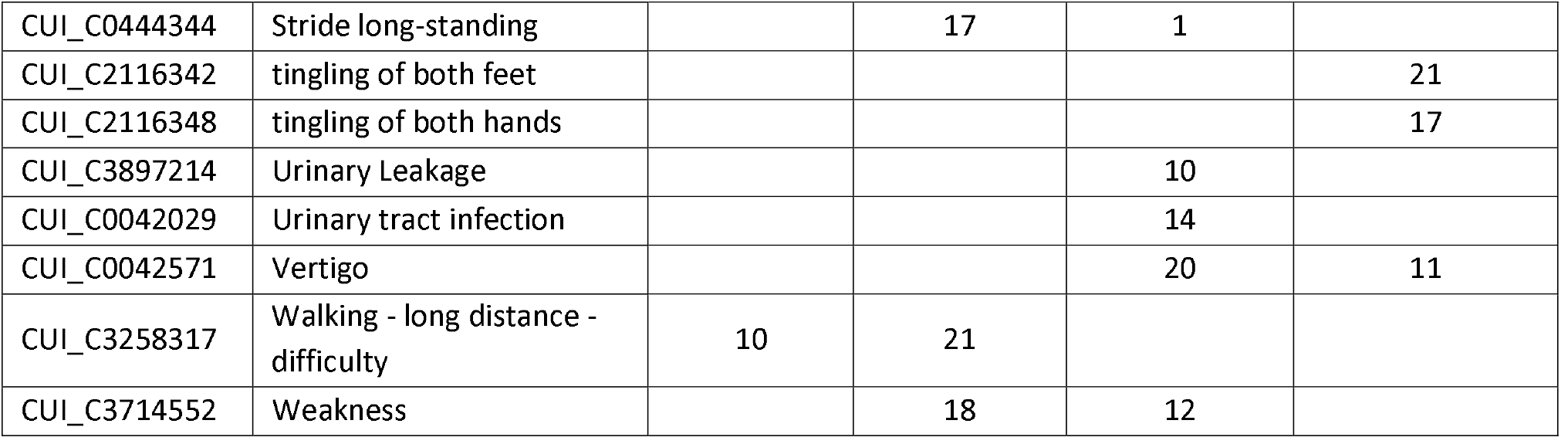
Top selected features (symptoms) and their coefficients ranking associated with each MS phenotype modifier.

The results demonstrated encouraging interpretability given lots of the selected features were persistently correlated to corresponding phenotype modifiers, such as “increased fatigue”, “mood swings”, “numbness of foot”, etc. However, the clinical relevance seems still incomplete with this method. For example, the highest ranked symptom associated with the “Active” modifier is not considered a widely common MS symptom. “Alopecia” is not a sign of MS but an adverse effect of several disease-modifying MS therapies. There are other seemingly unrelated symptom terms could be explained by an association with some of the more potent MS treatments, which may be used when a patient has activity or disease worsening. For example, Gilenya (fingolimod) and Tecfidera (dimethyl fumarate) are FDA approved oral MS disease modifying therapies that can be linked to the concepts “Macular retinal edema” (adverse effect of fingolimod) and “Psoriasis” (dimethyl fumarate has a similar structure to treatments for psoriasis).

## 4. Discussion

The integrated method of NLP and shallow machine learning models in this study extracts largely relevant clinical information from progress notes and predicts MS phenotype modifiers with satisfactory performance, encouraging portability and interpretability.

The progress notes in the EMR include a wealth of information relating to disease activity and progression. Previous studies showed that there was no clear difference in case-detection algorithm accuracy between rule-based and machine learning methods of extraction [15]. However, the large variations in local context in the clinical notes (e.g., wording, documentation habits, note structure, etc.) demand excessive efforts in forming those rules and make the rule-based approaches less feasible and portable. The modern text classification methods that utilize deep neural network models do not require heavy annotation, but are often regarded as black-boxes and criticized by the lack of interpretability since these models cannot provide meaningful interpretation on how a certain prediction is made [16]. The method used in this study did not require hand-crafted rules or custom developed dictionary to determine phenotype modifiers. Instead, it leveraged the local context in progress notes by analyzing the note structure and used advanced NLP technologies to detect the evidence of disease activity and progression, which were used to predict phenotype modifiers with shallow machine learning method. These advantages made this method applicable, although not in plug-and-play manner, in other data environment with consideration of its local context during the deployment.

In other studies of text classification using machine learning based NLP in building phenotype systems, distributed word representation modeling (e.g., word2vec) was commonly used to process raw texts for deep learning models, including convolutional neural network (CNN), recurrent neural networks (RNNs), long short-term memory (LSTM), etc. [17, 18]. The deep learning architecture provides good generalizability and reduce annotation complexity for clinical domain experts [11], a major drawback is the lack of interpretability. It can be difficult to understand how the predictions were made from the input features even though they can train a classifier with good performance [19]. Furthermore, unsupervised clustering models are getting increasingly adapted in biomedical domains, which helps to discover novel phenotypes. The meaningful interpretations are still challenging. The method in this study generally demonstrated appropriate clinical relevance, thus enhanced the interpretability of the prediction model.

Analyzing the inclusion/exclusion of clinical trials for MS showed that 40% of MS trials require characteristics of disease activity and progression only, rather than the base phenotypes. In this context, the methods demonstrated in this paper could be used to study the efficacy of screening patients for clinical trials using a machine learning based method versus manual selection. This method could also be utilized to study whether patients are receiving disease modifying therapy consistent with their MS characteristics. It can be very challenging to identify when patients transition from relapsing remitting to secondary progressive MS. One exciting potential application of this method is to see whether it can help categorize MS patients in a way that allows for early identification of this transition.

In addition to the positive aspects of this study, there are some limitations. It was conducted exclusively within the MS clinic at the University of Rochester. The MS patient population at UR has similar characteristics to the general MS population, but there may still be site-specific nuances to study cohort. Additionally, the volume of patient data in this study is relatively low, largely due to the time-consuming nature of labeling patient notes. A small study cohort has negative impacts on the overall performance of both NLP model and machine learning classification. It also leads to another major limitation in that a small study corpus likely has an imbalanced data issue not easily resolved. For example, the overall sensitivity and specificity of modifier “Progression” are higher than the others; this is very likely because the labels of progression are imbalanced, meaning the number positive labels (=Yes) are much lower than the number of negative labels (=No), compared with the positive to negative ratio in other labels of modifiers.

Local context in clinical notes in EMR imposes challenges in generalizability of information extraction (IE) methods involving NLP and/or rule-based approaches. Pre-analyzing the context and incorporating them in the overall design could help make the IE methods more portable, there may still be culture differences at each health care organization that influence the clinical note structure, documentation habits, language used that could have unexpected impacts on and further limits the generalizability of the IE method.

## 5. Conclusion

This study demonstrated that the integrated method of NLP with machine learning classification is capable of detecting evidence of disease activity and clinical progression from clinical notes in EMR. The classification algorithms are relatively simple but yield interpretable and largely clinically relevant features (symptoms and clinical conditions) that are persistently associated with disease activity and progression. Incorporation of a larger volume of labeled progress notes or involvement of additional clinical sites may further improve model performance and generalizability. Future applications for this work include facilitation of high throughput of MS clinical trial screening, refinement of disease modifying therapy utilization, and deeper analysis of the transition from relapsing to progressive MS phenotype.

## Data Availability

The data that support the findings of this study will be publicly available from Dryad.

